# Hydroxychloroquine (HCQ) reverses anti-PD-1 immune murine checkpoint blockade: TCF1 as a marker in humans for COVID-19 and HCQ therapy

**DOI:** 10.1101/2020.09.29.20193110

**Authors:** Janna Krueger, Francois Santinon, Alexandra Kazanova, Mark Issa, Bruno Larrivee, Catalin Milhalcioiu, Christopher E. Rudd

**Author notes:** To whom correspondence should be addressed- Christopher E. Rudd.

## Abstract

Coronavirus disease 2019 (COVID-19) pandemic caused by the severe acute respiratory syndrome coronavirus 2 (SARS-CoV-2) is a serious threat to global public health. Hydroxychloroquine (HCQ) and the antibiotic azithromycin (AZ) are still being used by thousands and numerous hospitals to treat COVID-19. In a related context, immunotherapy using checkpoint blockade (ICB) with antibodies such as anti-PD-1 has revolutionised cancer therapy. Given that cancer patients on ICB continue to be infected with SARS-CoV-2, an understanding of the effects of HCQ and AZ on the elimination of tumors by anti-PD-1 ICB is urgently needed. In this study, we report that HCQ alone, or in combination with AZ, at doses used to treat COVID-19 patients, reverses the therapeutic benefit of anti-PD-1 in controlling B16 melanoma tumor growth in mice. No deleterious effect was seen on untreated tumors, or in using AZ alone in anti-PD-1 immunotherapy. Mechanistically, HCQ and HCQ/AZ inhibited PD-L1 expression on tumor cells, while specifically targeting the anti-PD-1 induced increase in progenitor CD8^+^CD44^+^PD-1^+^TCF1^+^ tumor infiltrating T-cells (TILs) and the generation of CD8^+^CD44^+^PD-1^+^ effectors. Surprisingly, it also blocked the appearance of a subset of terminally exhausted CD8+ TILs. No effect was seen on the presence of CD4+ T-cells, FoxP3+ Tregs, thymic subsets, B-cells, antibody production, myeloid cells, or the vasculature of mice. Lastly, we identified TCF-1 expression in peripheral CD8+ T-cells from cancer or non-cancer human patients infected with SARs CoV2 as a marker for the effects of COVID-19 and HCQ on the immune system. This study indicates for the first time that HCQ and HCQ/AZ negatively impact the ability of anti-PD-1 checkpoint blockade to promote tumor rejection.

**Graphic Abstract:** 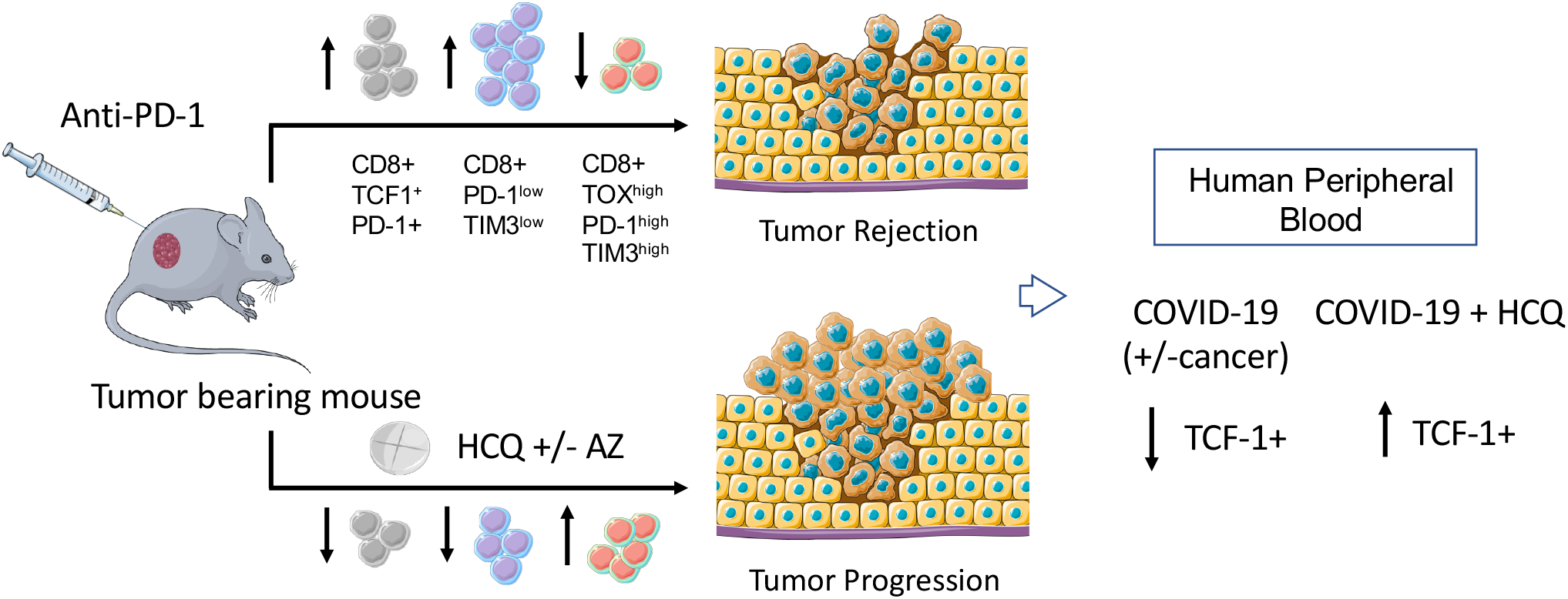

## Introduction

The outbreak of coronavirus disease 2019 (COVID-19) caused by the severe acute respiratory syndrome coronavirus 2 (SARS-CoV-2/2019-nCoV) poses a serious threat to global public health. The anti-malarial 4-aminoquinoline drugs chloroquine (CQ) and hydroxychloroquine (HCQ) as well as the antibiotic azithromycin (AZ) have gained much attention potential as potential therapies. The parent compound chloroquine (CQ) was originally reported to inhibit SARS-Cov1 coronavirus infection (Vincent et al., 2005) and *in vitro* studies have shown activity against SARS-CoV-2 (Liu et al., 2020; Wang et al., 2020). The exact mechanism of action of HQ and HCQ is unknown although these drugs increase the pH of endosomes that the virus uses for cell entry and can interfere with the glycosylation of the cellular receptor of SARS-CoV, angiotensin-converting enzyme 2 (ACE2) and associated gangliosides (Devaux et al., 2020). Other mechanisms have also been proposed (Quiros Roldan et al., 2020). CQ has a half-life of 20–60 days and can accumulate at higher levels in metabolically active tissues (Wong et al., 2020). Similarly, a macrolide antibiotic AZ can synergise with HCQ to block viral entry into cells and decrease viral replication (Andreani et al., 2020).

Despite this, the effectiveness of HCQ in treating COVID-19 has been highly contentious (Kupferschmidt and Cohen, 2020; Lenzer, 2020). An initial non-randomised trial showed remarkable efficacy in combination with AZ in clearing SARs-Cov2 (Gautret et al., 2020). Other studies have produced conflicting results (Chen et al., 2020a; Chen et al., 2020b; Gao et al., 2020; Geleris et al., 2020; Magagnoli et al., 2020; Mahevas et al., 2020; Molina et al., 2020; Rosenberg et al., 2020; Tang et al., 2020; Yu et al., 2020). Recently, the Henry Ford Covid-19 Task Force found that when controlled for COVID-19 risk factors, HCQ alone and in combination with AZ was associated with a reduction in mortality (Arshad et al., 2020). However, adverse cardiovascular effects may predispose certain patient groups to ventricular arrhythmias (Giudicessi et al., 2020; Ray et al., 2012). The US Federal Drug Administration (FDA) and the World Health Organisation (WHO) halted HCQ trials due to a lack of efficacy in reducing death rates of already hospitalised patients (https://covid19treatmentguidelines.nih.gov/therapeutic-options-under-investigation). However, concerns about side-effects were then influenced by the withdrawal of papers (Mehra et al., 2020a; Mehra et al., 2020b). The FDA has granted emergency use authorisation for HCQ in the treatment of a limited number of hospitalised cases but has cautioned against its use outside of a clinical trial or hospital setting. In this context, aside from the treatment of malaria, HCQ has also been used widely over the past decade at lower concentrations to treat auto-immune diseases such as systemic lupus erythematosus and rheumatoid arthritis where it can reduce inflammation (Thome et al., 2014; Thome et al., 2013).

Given this situation, HCQ is still being used by many countries and thousands of patients globally to treat COVID-19. There are more than 200 trials presently underway around the world on its impact either as a prophylactic or for treatment for COVID-19. The efficacy may vary with the dose used in the various studies. One recent study from the multi-center COVID-19 RISK and Treatments (CORIST) Collaboration, using 400-600 mg of HCQ day found that treatment was associated with reduced mortality (Di Castelnuovo et al., 2020). Another recent study from Belgium using similar doses also reported reduced mortality (Catteau et al., 2020). The doses and onset of treatment in these studies differed from the UK RECOVERY trial where 1600 mg on the first day and 800 mg on days 2-9 were given (Horby et al., 2020). Overall, there is an interest whether HCQ might be more effective if used early in infection, before the need for hospitalisation. A large COPCOV trial on the role of HCQ as a prophylactic for COVID-19 health workers was recently re-approved by the regulatory authorities in the United Kingdom (https://www.tropmedres.ac/covid-19/copcov). Funded by the COVID-19 Bill & Melinda Gates Foundation, Wellcome and Mastercard Therapeutics Accelerator grant, the COPCOV study will enroll over 40,000 frontline health care workers who have close contact with COVID-19 patients to determine whether chloroquine or hydroxychloroquine are effective in preventing COVID-19. Many countries continue to use HCQ to treat COVID-19 health workers who are suspected or confirmed of infection (Nina and Dash, 2020).

HCQ and CQ have been reported to have mixed and contradictory effects on the immune system. One report showed that CQ enhances human CD8+ T cell responses (Accapezzato et al., 2005), while another showed that HCQ inhibits CD4+ T-cell activation (Wu et al., 2017). HCQ is well known to inhibit autophagy (Bonam et al., 2020), may reduce the efficacy of antigen-presentation (Thome et al., 2014; Ziegler and Unanue, 1982) and decrease the production of proinflammatory Th17-relatedccytokines (Wozniacka et al., 2008). Another report showed that HCQ enhanced the function of suppressive regulatory T cells (Tregs) (Thome et al., 2013) while others have reported that HCQ increases the presence of Tregs and plasmacytoid dendritic cells together with a decrease in activated CD4+ T-cells and plasmacytoid dendritic cells in patients infected with the human immunodeficiency virus (HIV-1) (Piconi et al., 2011).

In this context, the past 15 years have witnessed a revolution in the application of immunotherapy for the treatment of cancer. Immune checkpoint blockade (ICB) uses monoclonal antibodies that block the binding of inhibitory receptors (IRs) on T cells to their natural ligands often expressed on cancer cells. The blockade of cytotoxic T-lymphocyte–associated antigen 4 (*CTLA*-*4*) and programmed death 1 (PD-1) or the PD-1 ligand, PD-L1 have achieved survival rates of 20-30% in treating cancers such as non-small cell lung carcinoma (NSCLC), melanoma, kidney, and bladder cancer (Page et al., 2013; Sharma et al., 2011). The tumor microenvironment (TME) can alter the make-up and function of TILs (Balkwill and Mantovani, 2001).

With the advent of the COVID-19 crisis, an increasing number of cancer patients on ICB are being infected with SARS-CoV2. This raises an important question as to the best therapeutic approach for COVID-19 patients, one that limits viral spread, while allowing for the benefit of checkpoint immunotherapy in promoting reactivity against tumor neo-antigens. This has been further complicated by reports that HCQ can reverse the drug sequestration in lysosomes (Li et al., 2018) and enhance chemo-sensitization in cancer patients (Li et al., 2018). Phase II trial studies showed that HCQ is effective in treating patients with recurrent oligometastatic prostate cancer by promoting cell death in cancer cells. In breast cancer, autophagy has been reported to be tumour-suppressive (Cianfanelli et al., 2015; Cicchini et al., 2014), while in other cases, can promote tumors (Karsli-Uzunbas et al., 2014; Yang et al., 2014). In a similar vein, AZ can inhibit primary antibody responses, and recall responses on bacterial infections (Fernandez et al., 2004). Although these previous studies emphasized effects on the gut microbiota (Routy et al., 2018; Vetizou et al., 2015), it is noteworthy that AZT and ciprofloxacin (CPX) also act as lipophilic weak bases where they affect intracellular organelles similar to CQ and HCQ (Poschet et al., 2020).

Given the urgent clinical context, there is a pressing need to assess the effects of both HCQ and AZ on the immune checkpoint response against cancer. In this study, we show that HCQ and AZ, alone or in combination, impaired the ability of anti-PD-1 therapy to promote the elimination of the B16 melanoma. We further show that HCQ or HCQ/AZ combined therapy selectively inhibits the appearance of self-renewing CD8+ PD-1+TCF1+ TILs and CD8+PD-1+TOX+ effector T-cells. We also identified TCF-1 expression as a marker in patient samples for the action of COVID-19 and HCQ. Our study indicates that HCQ negatively affects anti-PD-1 immune checkpoint blockade in the promotion of tumor rejection.

## Results

### HCQ and AZ block anti-PD-1 therapy

To address this issue, we assessed whether HCQ or AZ was beneficial, or harmful to tumor immunotherapy. Mice were implanted with B16-PD-L1 melanoma cells intradermally into C57B6/J mice 4 days before the injection of anti-PD-1 alone (200ug/mouse), or in conjunction with HCQ or AZ at days 4, 7, 11 and 14. Tumors were harvested on day 17. The dosing with HCQ and AZ was the same as used in clinical trials to treat SARS-CoV-2 infection (Gautret et al., 2020). **Fig. 1A** shows that the growth curve in which tumour volume increased from 100mm^3^ on day 10 to 610mm^3^ from day 16 (n=14). Anti-PD-1 reduced the growth of tumour as early as day 12 (i.e. 100 vs 210 mm^3^) until day 16 (i.e. 220 vs 610mm^3^). Importantly, HCQ impaired the anti-PD-1 reduction in tumor growth as seen on day 16 (i.e. 425mm^3^ for HCQ/anti-PD-1 vs. 210mm^3^ for anti-PD-1). Injection of AZ alone showed a trend in reducing the beneficial effect of anti-PD-1 (375mm^3^ vs 210mm^3^ for anti-PD-1), although this did not achieve statistical significance. The combination of HCQ and AZ reversed the benefits of anti-PD-1 to the same extent as HCQ alone (430mm^3^ vs 220mm^3^ for anti-PD-1) (also see spider graphs in **Figs. S1**,**2**). Interestingly, neither HCQ nor AZ affected tumor growth in the absence of anti-PD-1 (**Fig. 1B)**. Examples of control and treated tumours are shown in **Fig. 1C**. Neither drug treatment resulted in major loss of mouse weight (**Fig. 1D**), while scatter plot analysis showed a correlation between tumour volume and weight in reducing tumor size in response to anti-PD-1, HCQ and HCQ/AZ (p=0.025) (**Fig. S3**). HCQ did not directly affect the growth of *in vitro* cultured B16 cells (**Fig. S4**). These data showed that HCQ and HCQ/AZ impaired the ability of anti-PD-1 to promote tumor regression.

**Figure 1:**
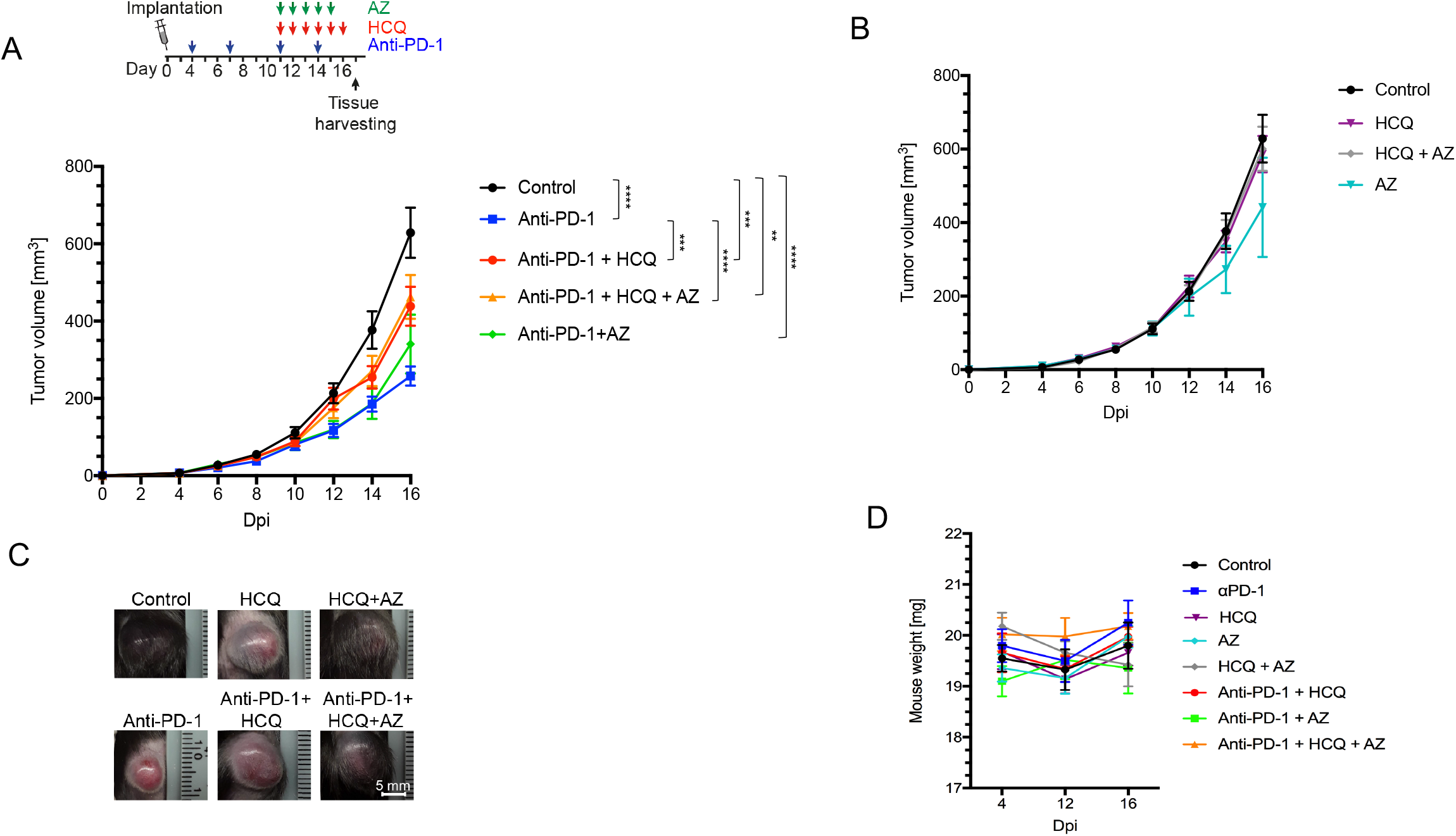
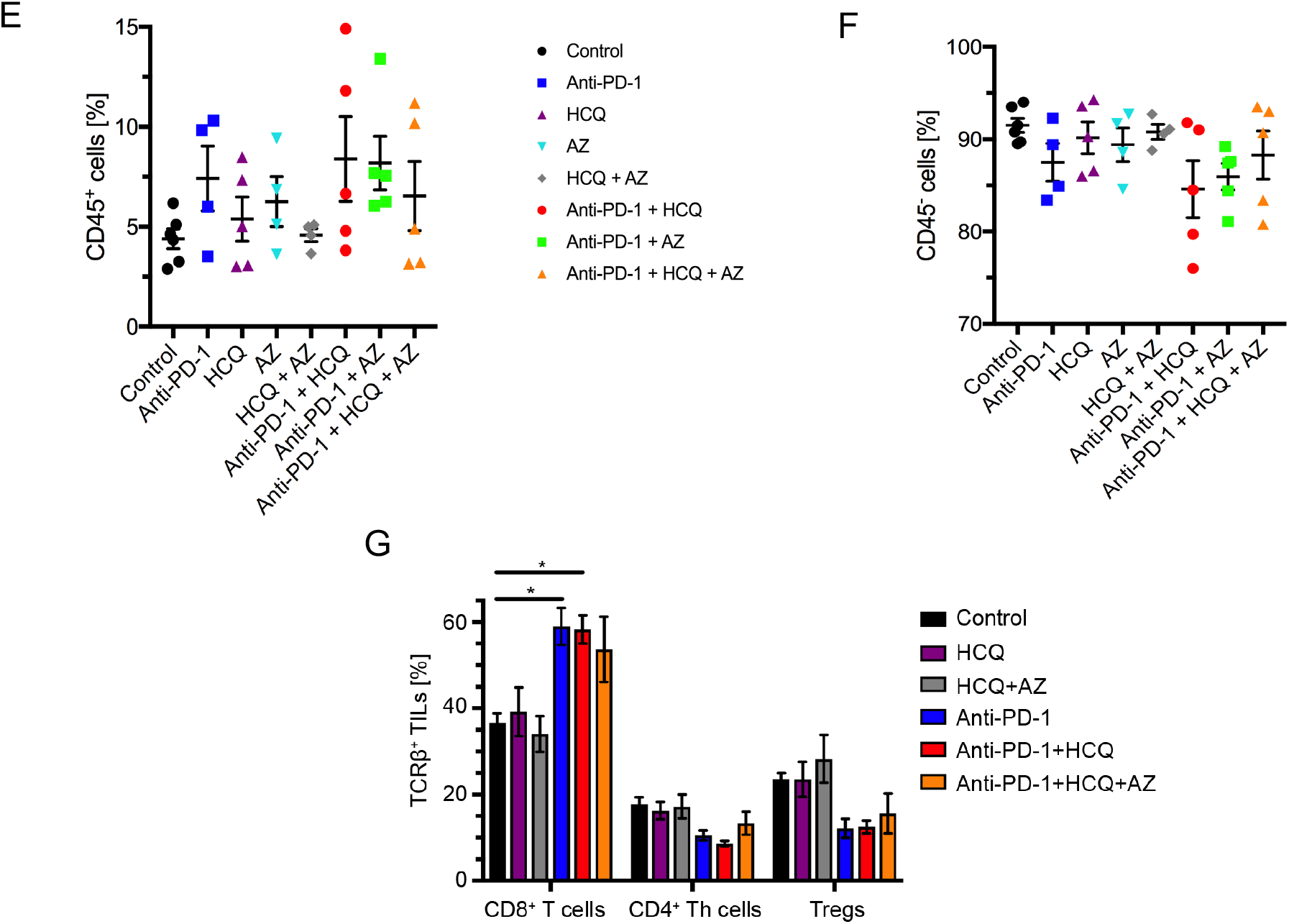
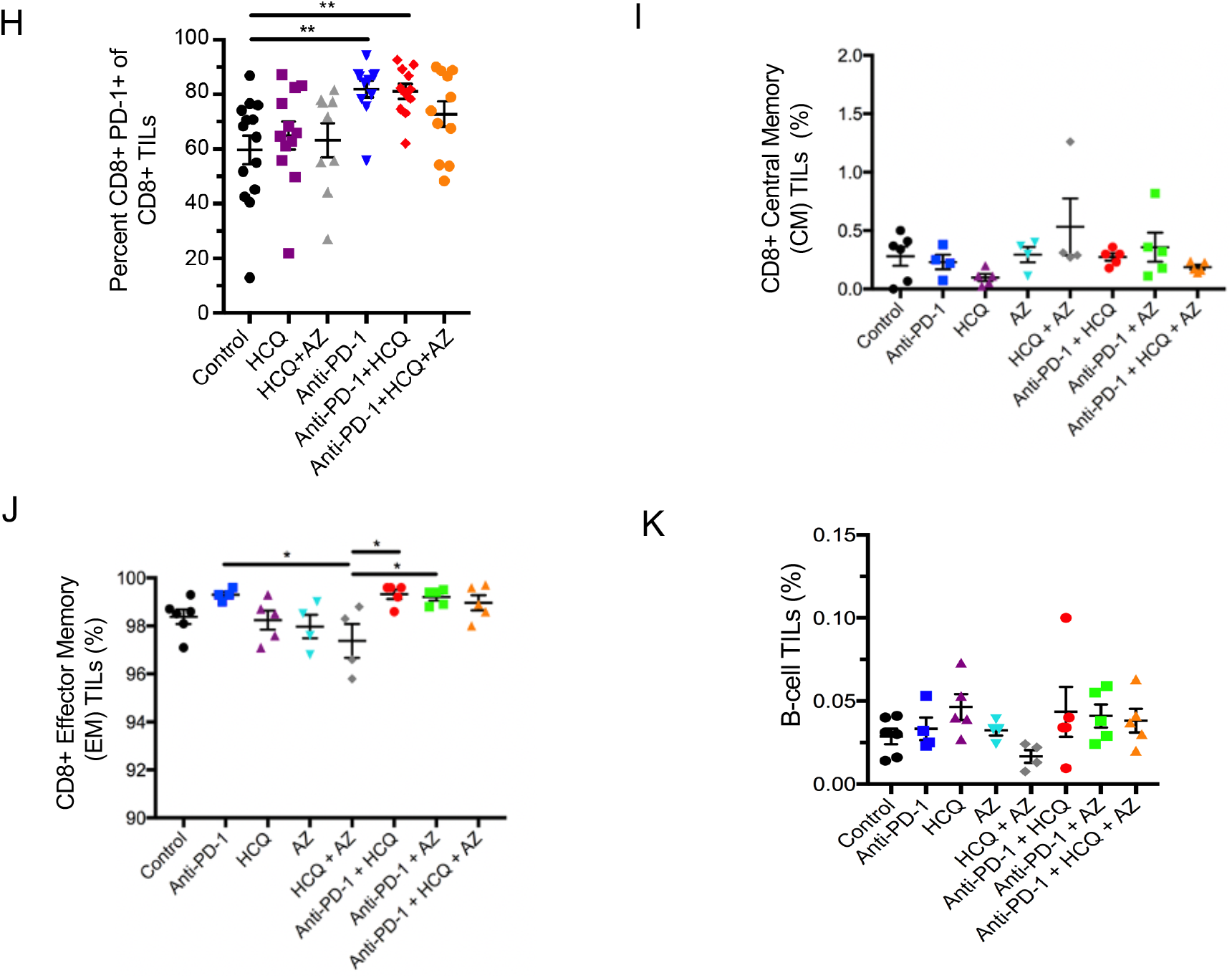
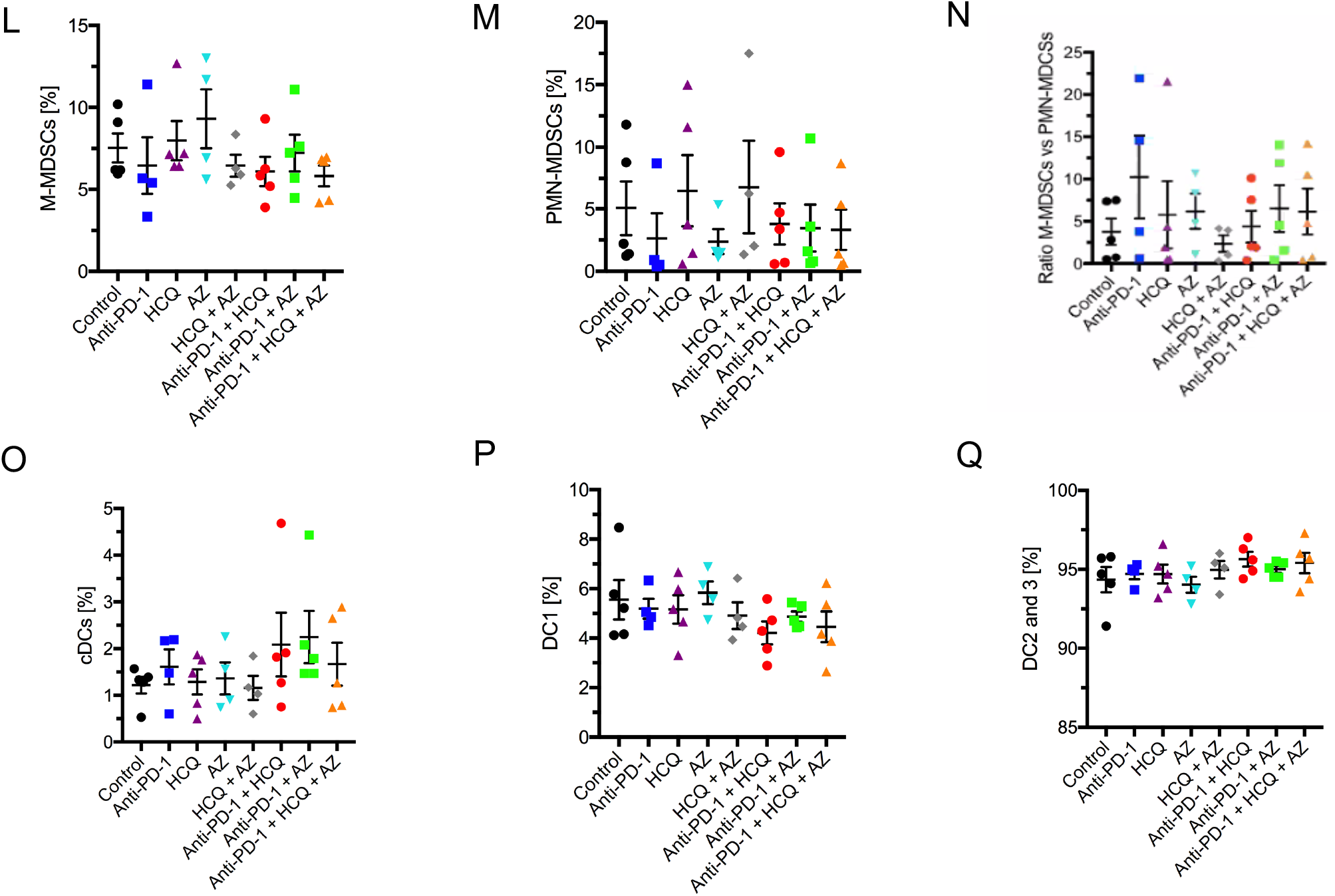
Hydroxychloroquine and azithromycin reverse partially immune checkpoint blockade in cancer therapy. C57BL/6 mice were implanted intradermally with B16-PD-L1 tumor cells. Following 1 week of anti-PD-1 treatment, HCQ with or without AZ were injected daily before tumors were harvested at day 17 post implantation. **Panel A:** Tumor volumes in response to anti-PD-1 plus HCQ and AZ. **Panel B:** Tumor volumes in mice treated only with HCQ and AZ. **Panel C:** Examples of tumors from mice treated with HCQ, AZ or anti-PD-1 plus HCQ and AZ. **Panel D:** Percentage of mouse weight loss over time. **Panel E:** Percentage of total immune cell infiltrate (CD45^+^ cells) **Panel F:** Percentage of CD45^-^ cells in tumors **Panel G:** Percentage of CD8^+^, CD4^+^ T helper and regulatory T cells (Tregs) of total TCRb^+^ cells in tumor- **Panel H:** Percentage of PD-1^+^ CD8^+^ TILs **Panel I:** The effect of therapy on CD8+ central memory TILs **Panel J:** The effect of therapy on CD8+ effector memory TILs **Panel K:** The effect of therapy on B-cell TILs. **Panel L:** The effect of therapy on the percentage representation of M-MDSC TILs. **Panel M:** The effect of therapy on the percentage representation of PMN-MDSC TILs. **Panel N:** Ratio of M-MDSC TILs relative to PMN-MDSC TILs. **Panel O:** The effect of therapy on the percentage representation of cDC TILs **Panel P:** The effect of therapy on the percentage representation of DC1 TILsc **Panel Q:** The effect of therapy on the percentage representation of DC2 and 3 TILs

We next examined the composition of TILs in tumors where anti-PD-1 showed a trend in increasing CD45+ tumour infiltrating lymphocytes (TILs) (**Fig. 1E**). Neither HCQ nor AZ affected this overall increase. Conversely, anti-PD-1 showed a trend in reducing the percentage of CD45-cells, although again, this was not statistically significant (**Fig. 1F**). In examining specific subsets, we showed that anti-PD-1 caused a significant increase in the overall presence of CD8+ TCRβ+ TILs, which was unaffected by HCQ or AZ (**Fig. 1G**). The same treatment showed a trend in decreasing the percentage of CD4+ T-cells and CD4+ FoxP3+ TILs. Neither AZ, nor HCQ affected this trend.

Within the CD8 TIL compartment, anti-PD-1 therapy increased the representation of CD8+CD44+PD-1+ cells (**Fig. 1H**), while the presence of HCQ and AZ interfered with the ability of anti-PD-1 to achieve statistical significance relative to untreated mice. By contrast, no differences were noted in CD8+ central memory TILs (**Fig. 1I**) or CD8+ effector memory T-cells (**Fig. 1J**), B cells (**Fig. 1K**).

In terms of myeloid cells, cancer-driven emergency myelopoiesis give rises to myeloid-derived suppressor cells (MDSCs) that express PD-1 (Rudd, 2020; Strauss et al., 2020). Anti-PD-1 therapy showed a trend in decreasing the presence of suppressive M-MDSCs (**Fig. 1L**) and PMN-MDSCs (**Fig. 1M**) as well as in increasing the ratio of M-MDSCs to PMN-MDSCs **(Fig. 1N**), as reported (Strauss et al., 2020). HCQ nor AZ had a statistically irrelevant effect on this trend. Similarly, anti-PD-1 showed a trend in increasing the presence of cDC TILs which was unaffected by HCQ or HCQ/AZ (**Fig. 1O**). No obvious effect of therapies on the presence of DC1 (**Fig. 1P**) or DC2 and 3 TILs was observed (**Fig. 1Q**).

We next examined the surface expression of PD-1, PD-L1 and CD80 receptors on different TILs (**Fig. 2**). Anti-PD-1 treatment showed a trend in increasing PD-1 expression on PMN-MDSCs **(Fig. 2A**). Neither HCQ nor AZ monotherapy in combination with anti-PD-1 affected this trend, although the combination of HCQ/AZ reduced expression to control levels. No effect on the expression of PD-L1 or CD80 was observed. Anti-PD-1 increased the percentage of PD-1 expressing PMN-MDSCs which was unaffected by co-injection of HCQ or AZ in mice (**Fig. 2B**). The percentage of PMN-MDSCs TILs expressing PD-L1 was unaffected by anti-PD-1 or anti-PD-1 with HCQ or AZ (**Fig. 2B**). Similarly, no effect was observed on the expression of class 2 major histocompatibility antigens (MHC) on B-cells (**Fig. 2C**) or cDCs (**Fig. 2D**).

**Figure 2:**
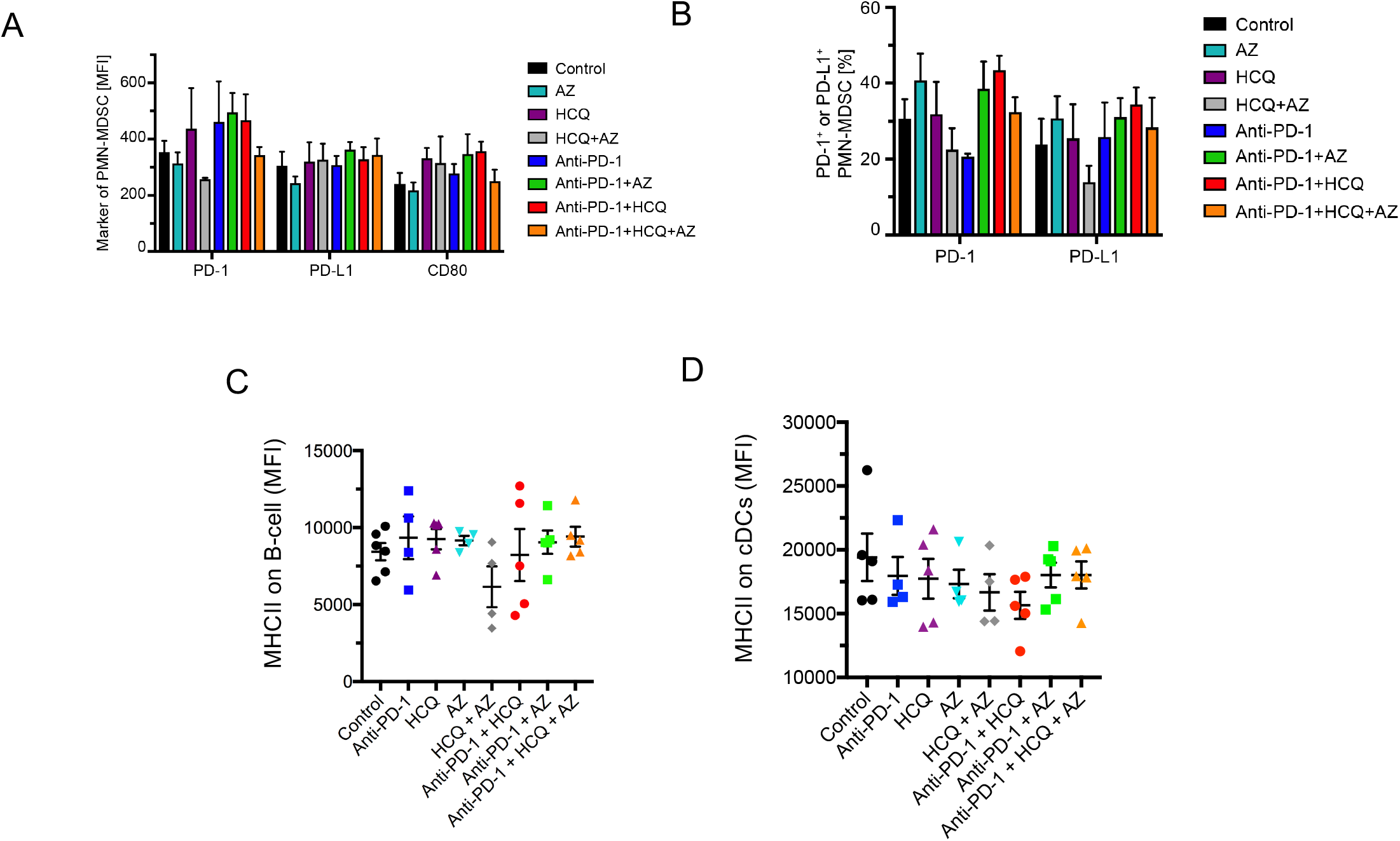
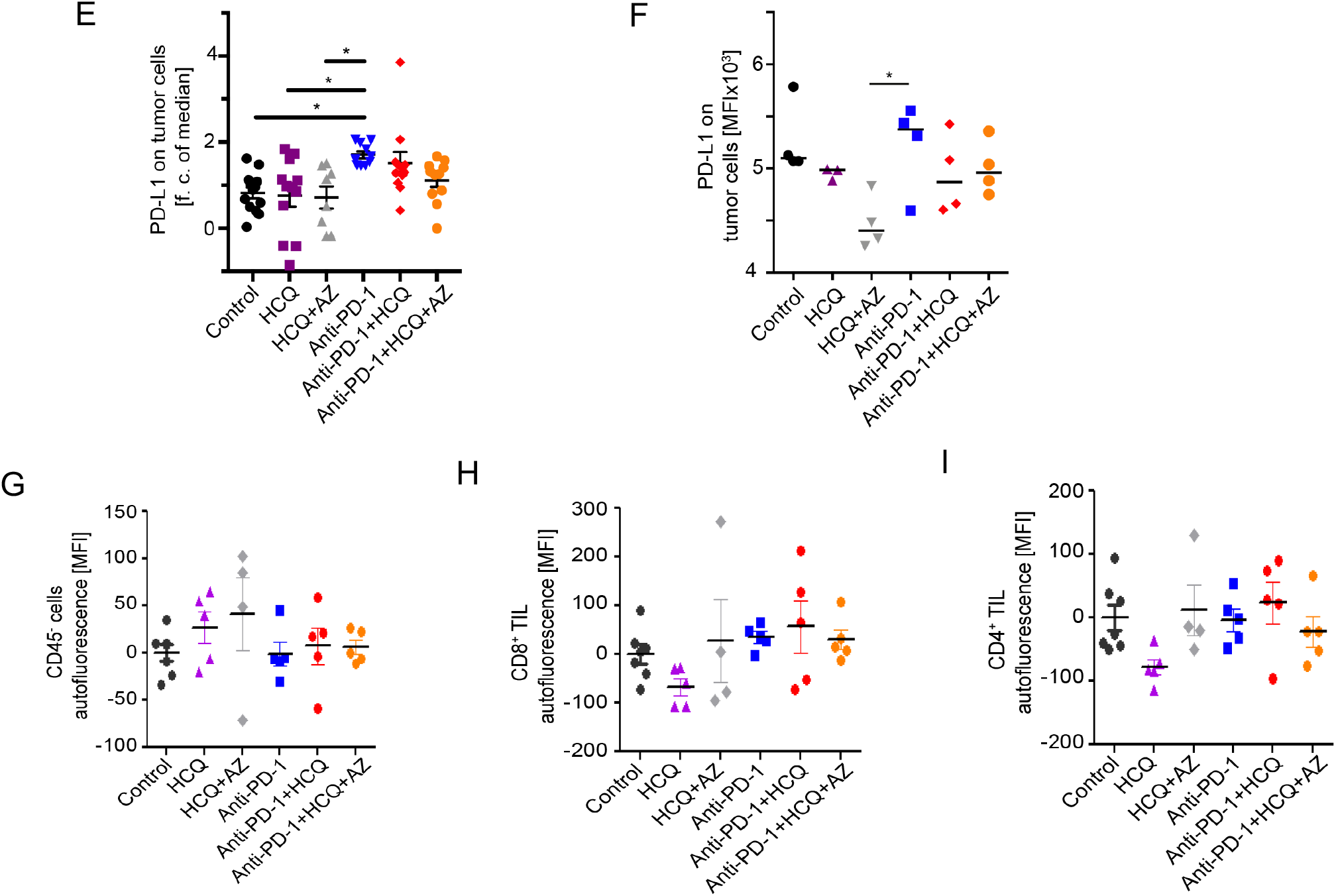
The effect of HCQ and AZ on the expression of PD1, PD-L1 and CD80 on TIL subsets. **Panel A:** Histogram showing the medium expression of PD1, PD-L1 and CD80 on PMN-MDSC TILs. **Panel B:** Histogram showing the percent positive PMN-MDSC TILs expressing PD1 and PD-L1. **Panel C:** The effect of therapy on MHC class II expression (MFI) on B-cells. **Panel D:** The effect of therapy on MHC class II expression (MFI) on cDCs. **Panel E:** The in vivo effect of HCQ and HCQ/AZ on the anti-PD-1 induced increase in PDL-1 expression on tumor cells (defined as CD45^-^ CD31^-^) **Panel F:** The effect of HCQ and HCQ/AZ on the anti-PD-1 induced increase in PDL-1 expression in vitro cultured B16 cells. **Panel G:** HCQ accumulation in non-immune cells (CD45^-^) TILs. **Panel H:** HCQ accumulation in CD8^+^ TILs. **Panel I:** HCQ accumulation in CD4^+^ TILs.

In contrast, anti-PD-1 increased the expression of PD-L1 on the surface of CD45-CD31-, SSC+ and FSC+ B16 tumor cells and this effect was inhibited by the co-expression of HCQ and HCQ/AZ (**Fig. 2E**). Similarly, when cultured *in vitro*, anti-PD-1 increased the expression of PD-L1 on B16 cells (**Fig. 2F**). Co-incubation with different concentrations of HCQ and HCQ/AZ inhibited the increase. These data showed that HCQ and HCQ/AZ could act in *in vitro* and *in vivo* to reduce an increase in PD-L1 expression on tumor cells that was induced by anti-PD1. No effect on the expression of other key ligands on myeloid, B-cells or on the B16 melanoma cells was observed.

Likewise, we did not observe changes in the presence of IgM or IgG antibodies against PD-L1 in mice treated with anti-PD1, alone, or in combination with HCQ/AZ (**Fig. S4**). Index value was calculated by normalizing B16-IgG and B16-IgM MFI of the sample to the average of tumor-free mice (**Fig. S4D-F**). We also did not observe effects on the composition of thymic subsets (**Fig. S5**). Lastly, as a control, we monitored the localisation of HCQ in cellular compartments using autofluorescence (Sauer et al., 2019). Autofluorescence was observed in the CD45-fraction which was mostly comprised of B16 cancer cells (**Fig. 2G**) as well as in the CD8+ and CD4+ TIL population (**Figs. 2H and I**, respectively).

### HCQ and AZ selectively inhibits anti-PD-1 induction of progenitor CD8+PD-1+TCF1+ TILs

Given the effect of HCQ and AZ on CD8+ TILs, we next used viSNE and Cytobank analysis to interrogate the composition of this subset (**Fig. 3**). viSNE can visually define groupings of immune cells by simultaneously combining multiple markers (Amir el et al., 2013). The CD8 compartment could be separated into different areas by staining with anti-PD-1 and anti-TCF-1 (**Fig. 3A**). One island of CD8+ cells showed moderate to high levels of PD-1 and CD44 expression (island (i)), while a separate grouping of cells expressed primarily TCF-1 but no PD-1 (island (ii)). A grouping of cells between islands (i) and (ii) showed moderate levels of PD-1 and TCF-1 co-expression (island (iii)). TCF-1 defines progenitor CD8+ T-cells (Raghu et al., 2019; Weber et al., 2011) which give rise to effector CD8+ T-cells (Page et al., 2013; Rudd, 2020).

**Figure 3:**
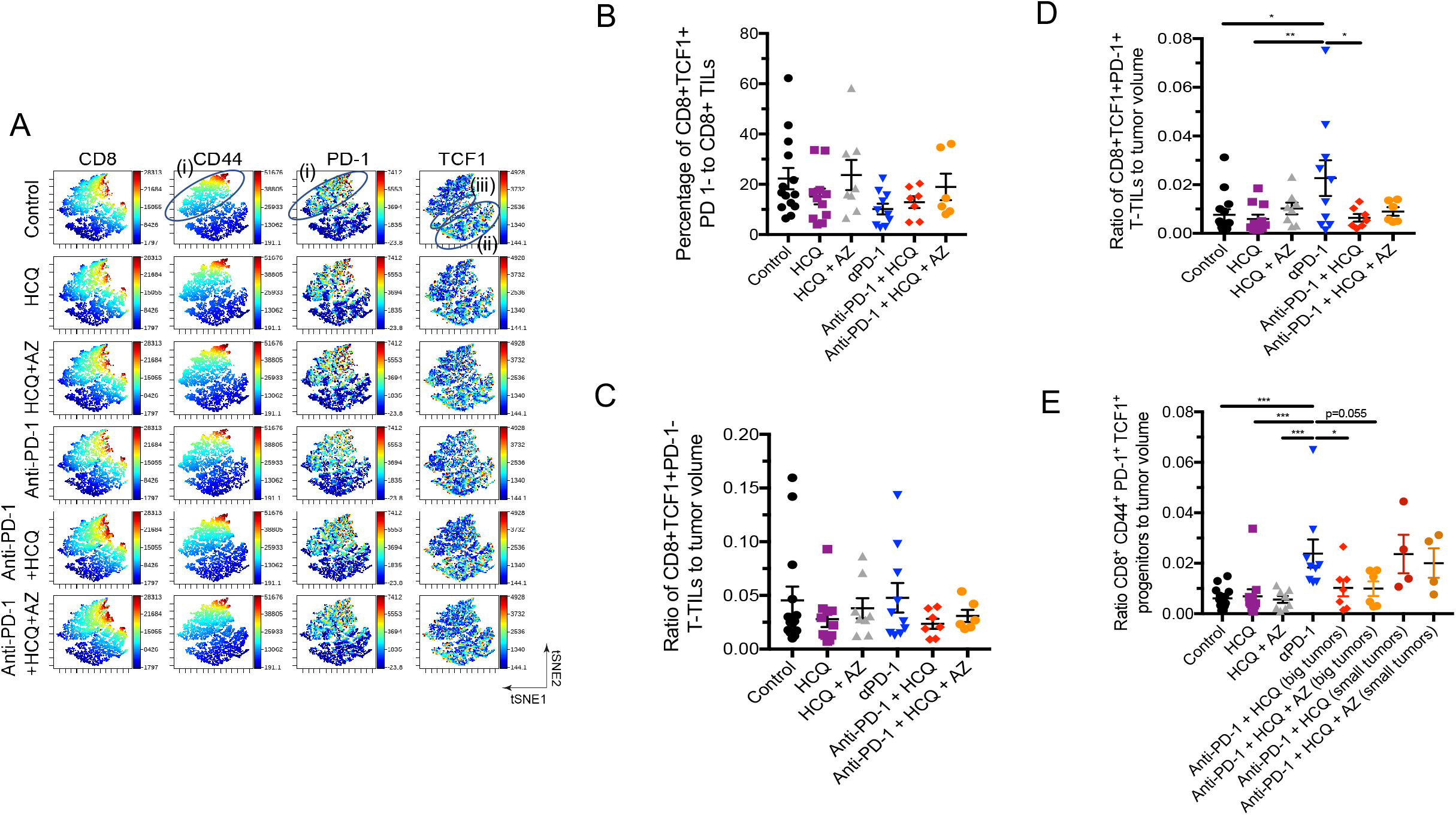
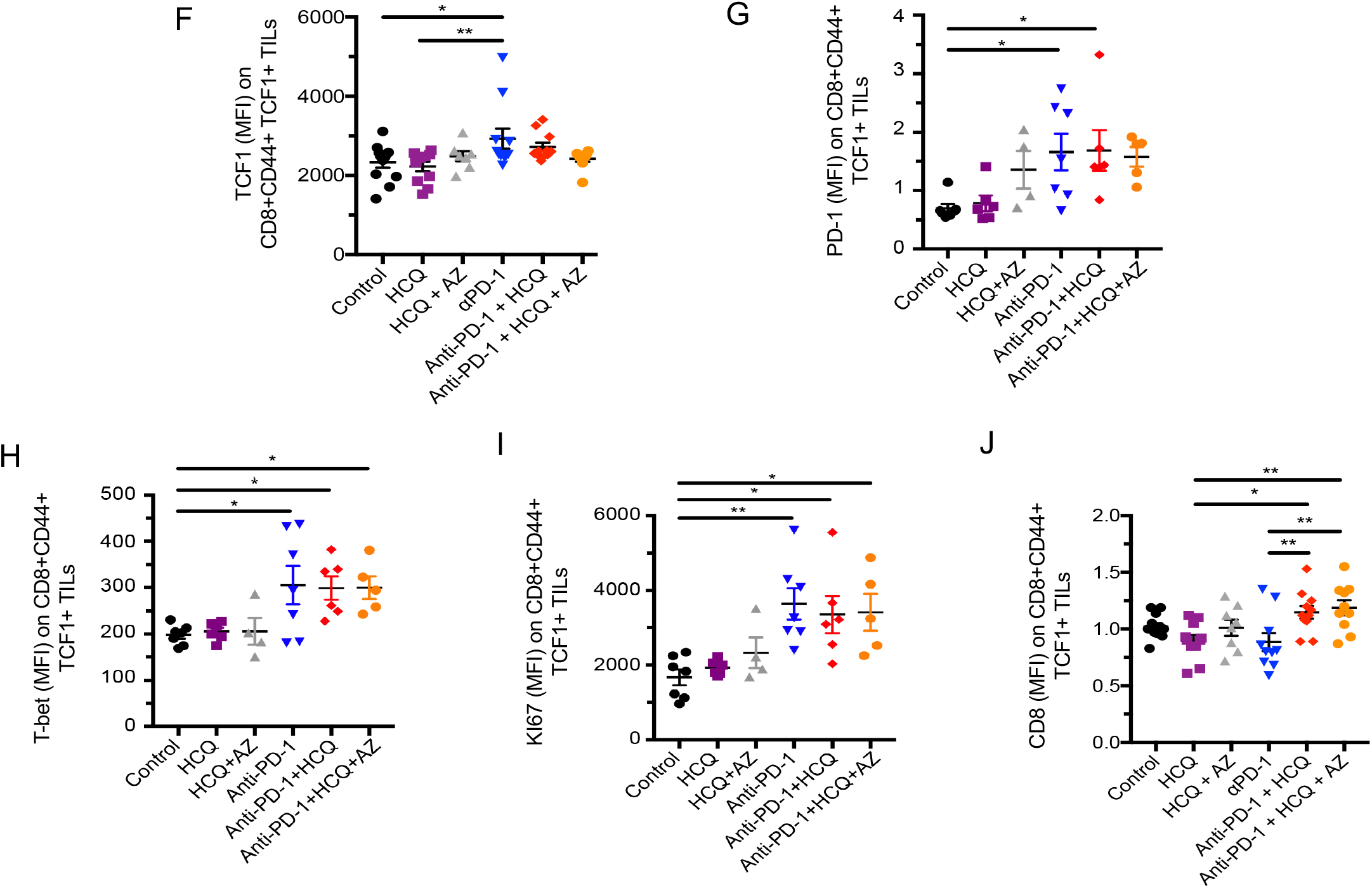
The effect of HCQ and AZ on the presence of CD8^+^TCF1^+^ progenitor TILs. Mice were implanted with B16 tumours as described in Figure 1. **Panel A:** viSNE profiles of total CD8^+^ TILs. The figure shows the presence of the upper cluster of TILs expressing moderate to high levels of CD8, PD-1 and CD44 (island (i)), a second island (ii) expressing moderate levels of TCF-1^+^ with no PD-1; and island (iii) that co-expresses PD-1^+^ and TCF-1^+^. **Panel B**: Percentage of CD8+ TCF-1+ PD-1-TILs as a percent of total CD8+ TILs. **Panel C:** Ratio of CD8+ CD44+TCF-1+ PD-1-TILs relative to tumor volume. **Panel D:** Ratio of CD8+ TCF-1+ PD-1+ TILs relative to tumor volume. **Panel E:** Ratio of CD8+ CD44+ PD-1+TCF-1+ progenitor TILs relative to tumor volume. A comparison of tumor volumes in mice where HCQ blocked the response to anti-PD-1 (big tumors) and mice where HCQ failed to block the response to anti-PD-1 (small tumors). **Panel F:** MFI of TCF-1 expression CD8+ CD44+TCF-1+ PD-1+ TILs in response to anti-PD-1 alone or in combination with HCQ or HCQ/AZ. **Panel G:** MFI of PD-1 expression CD8+ CD44+TCF-1+ PD-1+ TILs in response to anti-PD-1 alone or in combination with HCQ or HCQ/AZ. **Panel H:** MFI of T-bet expression CD8+ CD44+TCF-1+ PD-1+ TILs in response to anti-PD-1 alone or in combination with HCQ or HCQ/AZ. **Panel I:** MFI of Ki67 expression CD8+ CD44+TCF-1+ PD-1+ TILs in response to anti-PD-1 alone or in combination with HCQ or HCQ/AZ. **Panel J:** MFI of CD8 expression CD8+ CD44+TCF-1+ PD-1+ TILs in response to anti-PD-1 alone or in combination with HCQ or HCQ/AZ.

We first assessed the presence of TCF-1+PD-1- and TCF-1+PD-1+ progenitor TILs in response to anti-PD-1 therapy, either alone or in conjunction with HCQ or AZ. Anti-PD-1 did not alter the presence of CD8+TCF-1+PD-1-cells relative to the overall CD8+ TIL population (**Fig. 3B**) or relative to the tumor volume (**Fig. 3C**). HCQ and HCQ/AZ also had no effect on the presence of this subset of TILs. By contrast, anti-PD-1 consistently increased the presence of CD8+TCF1+PD-1+ as measured relative to tumor volume **(Fig. 3D)**. Significantly, HCQ and HCQ/AZ inhibited this event. Similarly, anti-PD-1 therapy increased the presence of a subset of TCF1+PD-1+TILs expressing the receptor CD44 **(Fig. 3E)**. CD44 is a marker for antigen (Ag)-experienced, effector and memory T cells (Baaten et al., 2012). In this instance, we also compared the effects of HCQ and HCQ/AZ from tumors of responsive and non-responsive mice. Mice which were responsive to the effects of HCQ and HCQ/AZ had larger tumors (i.e. big), unaffected by anti-PD-1. By contrast, mice which were non-responsive to the effects of HCQ and HCQ/AZ had smaller tumors (i.e. small) similar in size to those seen with anti-PD-1 therapy alone. Tumors from responsive mice showed a clear inhibition of the presence of CD8+TCF-1+PD-1+ progenitors, while tumors where from HCQ non-responsive mice showed an increase in the presence of progenitors similar to that seen with anti-PD-1 therapy.

In terms of receptor expression, dot plot profiles showed that anti-PD-1 increased the MFI for TCF-1 expression on CD8+CD44+TCF1+ TILs (**Fig. 3F**). Co-therapy with HCQ and HCQ/AZ had marginal but significant effects on preventing anti-PD-1 from achieving statistical significance relative to untreated control mice. These data showed that anti-PD-1 increased both the numbers of CD8+CD44+TCF1+ cells as well as the level of TCF-1 expression on these cells and that these effects were inhibited by both HCQ and HCQ/AZ. Less obvious effects of HCQ on anti-PD-1 induced increases in PD-1 (**Fig. 3G**), the transcription factor T-bet (**Fig. 3H**) or the activation marker Ki67 were seen (**Fig. 3I**). Interestingly, HCQ and HCQ/AZ increased the expression CD8 when treatment was combined with anti-PD-1 (**Fig. 3J)**.

Overall, these data showed that HCQ has a specific effect in inhibiting the ability of anti-PD-1 immunotherapy to increase the generation of CD8+PD-1+TCF1+ and CD8+CD44+ PD-1+TCF1+ progenitor TILs relative to tumor volume.

### HCQ and AZ inhibited the ability of anti-PD-1 to induce CD8+ effector TILs

We next assessed the effects of HCQ and AZ on the generation of CD8+ effector TILs, which do not express TCF-1, but are derived from progenitors (Miller et al., 2019). viSNE analysis showed the patterns of cells stained with anti-CD8, CD44, PD-1, TOX and TIM-3 (**Fig. 4A**). 5 different clusters could be identified based on levels of receptor expression (**Fig. 4B**). This included TCF1-CD8+ cells with low-intermediate levels of PD1, TOX, TIM-3 (i.e. cluster 2: PD1+TOX+TIM-3+) as well as cells with higher levels of PD1, TOX, TIM-3 expression (i.e. clusters 3-5). Cells with the higher levels of PD-1 and TIM-3 expression classically correspond to exhausted T-cells (Jin et al., 2010; Wherry, 2011). The status of cells with lower levels of these inhibitory receptors is unclear, but most likely corresponds to activated functional CD8+ T-cells, which will eventually become exhausted, if exposed to repeated, ongoing antigenic stimulation.

**Figure 4:**
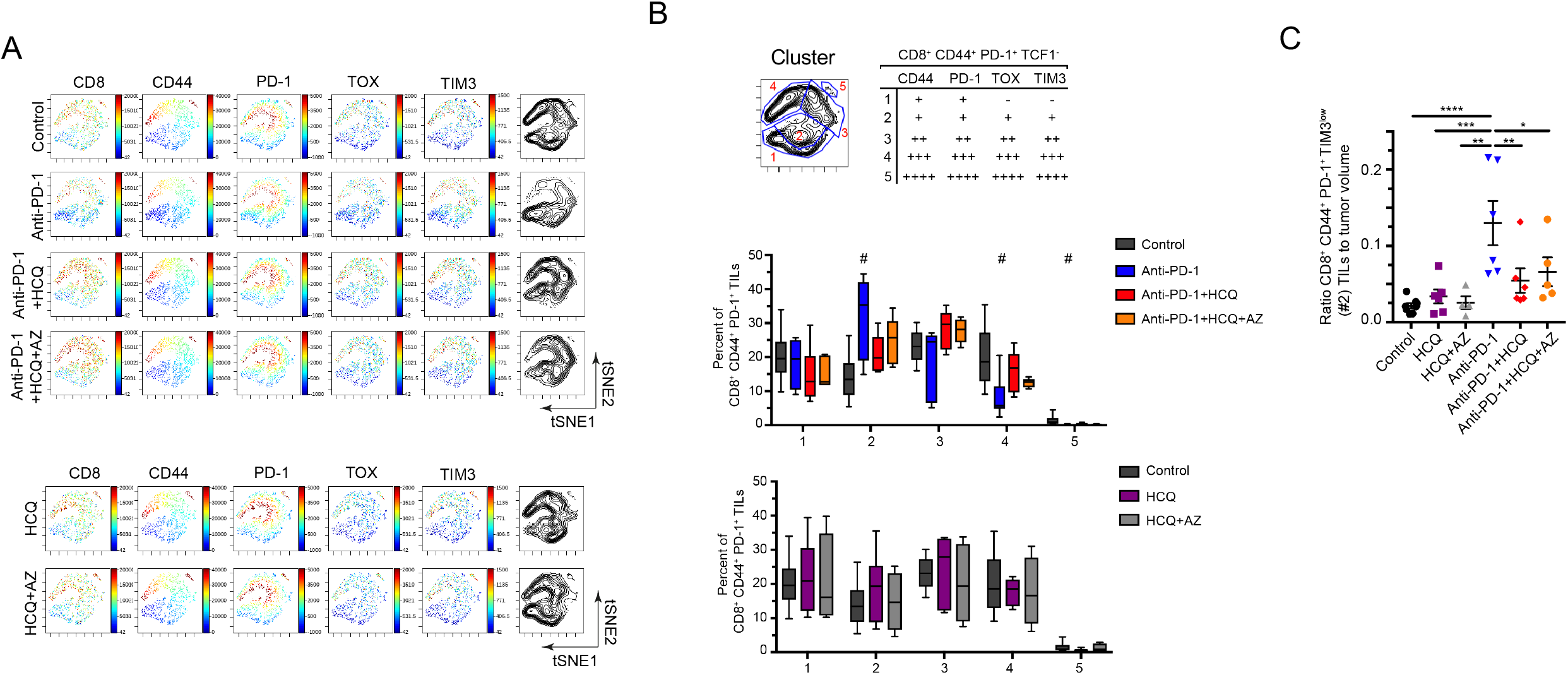
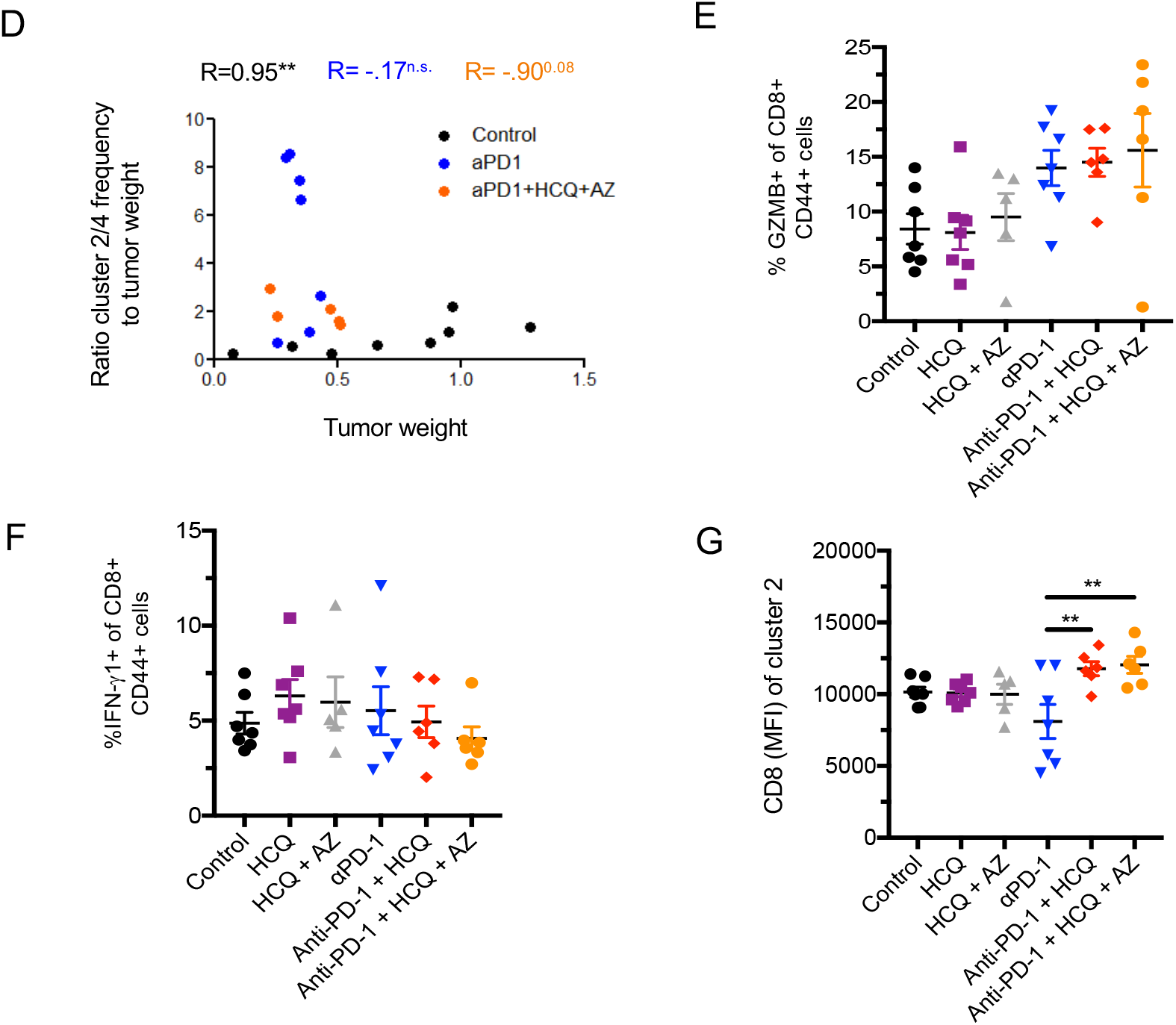
The expansion of specific CD8^+^TCF-1-CD44^+^ PD-1+ subsets in response to anti-PD-1 is inhibited by HCQ and AZ. **Panel A:** viSNE analysis of CD8, CD44, PD-1, TOX and TIM-3 expression of CD8^+^ CD44^+^ PD-1^+^ TILs. TCF1+ subsets have been removed **Panel B:** tSNE subdivision into different clusters based on expression levels of CD44, PD-1, TOX and TIM3. Upper panel: Table shows how clusters were grouped together. Middle panel: Histogram showing changes in the representation of different clusters in response to anti-PD-1 alone or in combination with HCQ and AZ. Lower panel: Histogram showing changes in the representation of different clusters in response HCQ and HCQ/AZ. **Panel C:** Ratio of cluster 2 relative to tumor volume. Anti-PD-1 induced an increase in CD8+ effectors which was inhibited by HCQ and HCQ/AZ. **Panel D:** Figure showing the ratio of cluster2/4 to tumor weight. **Panel E:** Percent of GZMB^+^ cells within the CD8^+^ CD44^+^ PD-1^+^ TIL population. **Panel F:** Percent IFNγ1+ cells within the CD8^+^ CD44^+^ PD-1^+^ TIL population. **Panel G:** CD8 expression (MFI) on cluster 2 TIL.

From this, we observed that anti-PD-1 induced a clear increase in TCF1-CD8+ cluster 2 expressing low levels of PD1, TOX, TIM-3 (i.e. PD1+TOX+TIM-3+) (upper table and middle panel). Significantly, HCQ and HCQ/AZ inhibited the increase in the presence of this population of cells. By contrast, no effect was seen on the presence of this population in anti-PD-1 untreated mice (lower panel). Dot plot analysis confirmed that anti-PD-1 increased the ratio of CD8+CD44+PD1+TOX+TIM-3^low^ TILs (cluster 2) relative to tumor volume and that this increase was blocked by the presence of HCQ or HCQ/AZ (**Fig. 4C**). These data showed that HCQ inhibited the expansion of CD8+ TILs induced by anti-PD-1 therapy, expressing low levels of the activation markers PD-1 and TIM-3, either due to the inhibition of the presence of TCF-1+PD-1+ progenitor T-cells and/or due to direct effects on the PD1+TOX+TIM-3+ cells themselves.

At the same time, anti-PD-1 decreased the presence of CD8+CD44+ T-cells with higher levels of PD-1, TIM-3 and TOX-3 corresponding to clusters 3-5 (**Fig. 4B**). In the case of clusters 4 and 5, the decrease was statistically significant. HCQ and HCQ/AZ treatment inhibited the anti-PD-1 induced reduction in the presence of exhausted T-cells as most clearly seen in cluster 4. Unfortunately, as in other studies (Wei et al., 2017), due to small numbers of recovered TILs, it was not possible to assess directly whether these subsets of cells were functionally non-responsive or dysfunctional.

Importantly, an analysis of the cluster 2 (effector-like) to cluster 4 (terminal exhaustion) frequency ratio relative to tumor weight further showed that shift to cluster 2 in response to anti-PD-1 and its inhibition by HCQ/AZ was correlated with the loss of tumor control (**Fig. 4D**).

Lastly, we observed that anti-PD-1 showed a trend in increasing the percentage of CD8+CD44+ T-cells expressing GZMB and this trend was unaffected by HCQ or HCQ/AZ (**Fig. 4E**). Anti-PD-1 also showed a trend in increasing the percentage of CD8+CD44+ T-cells expressing IFNγ1 which was unaffected by HCQ or HCQ/AZ (**Fig. 4F**). Lastly, as was seen in the progenitor population, HCQ and HCQ/AZ had an usual effect on the expression of CD8 where it cooperated with anti-PD-1 to significantly increase the MFI of CD8 expression on the CD8+ effector-like (cluster 2) T-cells (**Fig. 4G**).

Overall, these results showed that HCQ and HCQ/AZ interfered with the ability of anti-PD-1 to increase CD8+effector-like TILs while at the same time prevented CD8+ TILs from acquiring a phenotype indicative of T-cell exhaustion. A general ability of HCQ to inhibit T-cell activation would be consistent with this phenotype were it both inhibits the generation of functional CD8+ effectors while also preventing their progression into a more exhausted phenotype.

### HCQ and AZ did not affect antibody and vascular endothelial cells

Aside from the immune system, we also examined the vasculature in mice (**Fig. 5**). To evaluate potential alterations in the morphological features of blood vessels, we examined retinal and brain cortical vasculature in treated mice. Whole mounts of retinal vasculature showed no changes in retinal vessel morphology or branching between groups. (**Fig. 5A**). Further, an examination of the pial cerebral vessels, cortical vessels, vessel bifurcation and length showed no differences (**Fig. 5B**). Images of retinal and brain cortical vasculature are shown in **Fig. 5C**. These data indicate that neither anti-PD-1 nor HCQ or AZ had any detectable effects on the vasculature in mice. Furthermore, we found no effect of treatments on the overall branching structure in the cortical vasculature. These results show that the topological structure of the retinal and brain cortical vascular networks was not affected by the treatments.

**Figure 5:**
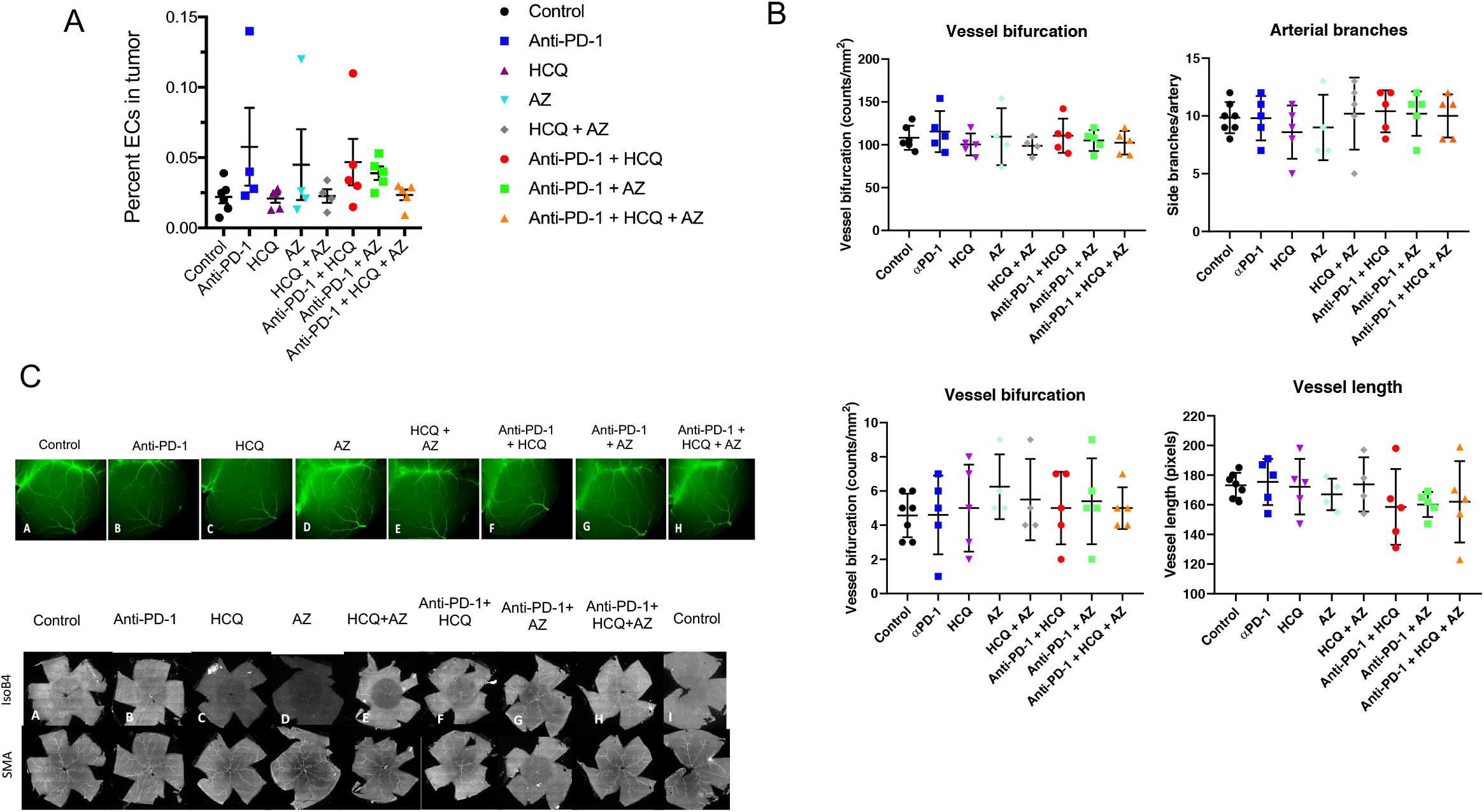
HCQ did not affect the vasculature. **Panel A:** Frequency of endothelial cells (ECs) relative to viable cells in the tumor. **Panel B:** Whole mount images of cerebral and retinal vessels **Panel C:** Quantification of vessel bifurcation, arterial branches and vessel length in response to different treatments of mice.

### TCF-1 expression on peripheral T-cells is a marker for COVID-19 and HCQ effects

Given the continuing widespread global use of HCQ in treating patients who are infected with SAR-CoV-2, we attempted to identify a phenotypic marker in human peripheral blood for the effects of COVID-19 and HCQ on the immune system (**Fig. 6**). Peripheral T-cells from the blood of cancer and non-cancer COVID-19 patients treated with HCQ were analyzed. viSNE analysis identified three separate clusters of CD8+ human T-cells, one island (i) staining with PD-1 (upper panel), Tim-3 (upper middle) and GZMB (lower panel) (**Fig. 6A**, upper circles), and a two other distinct islands (lower circles ii and iii) which stained clearly with anti-TCF-1 (lower middle panel). Images are a compilation of data from >20 patients. TCF-1+ clusters (ii) and (iii) could be distinguished on the basis of the expression of Ki67 and CCR7 which preferentially stained island (iii) (**Fig. 6B)**. Ki67 is an activation marker while CCR7 mediates the homing of T cells to secondary lymphoid organs via high endothelial venules (HEV) and can act as a marker for memory T-cells (Sharma et al., 2015)

**Figure 6.**
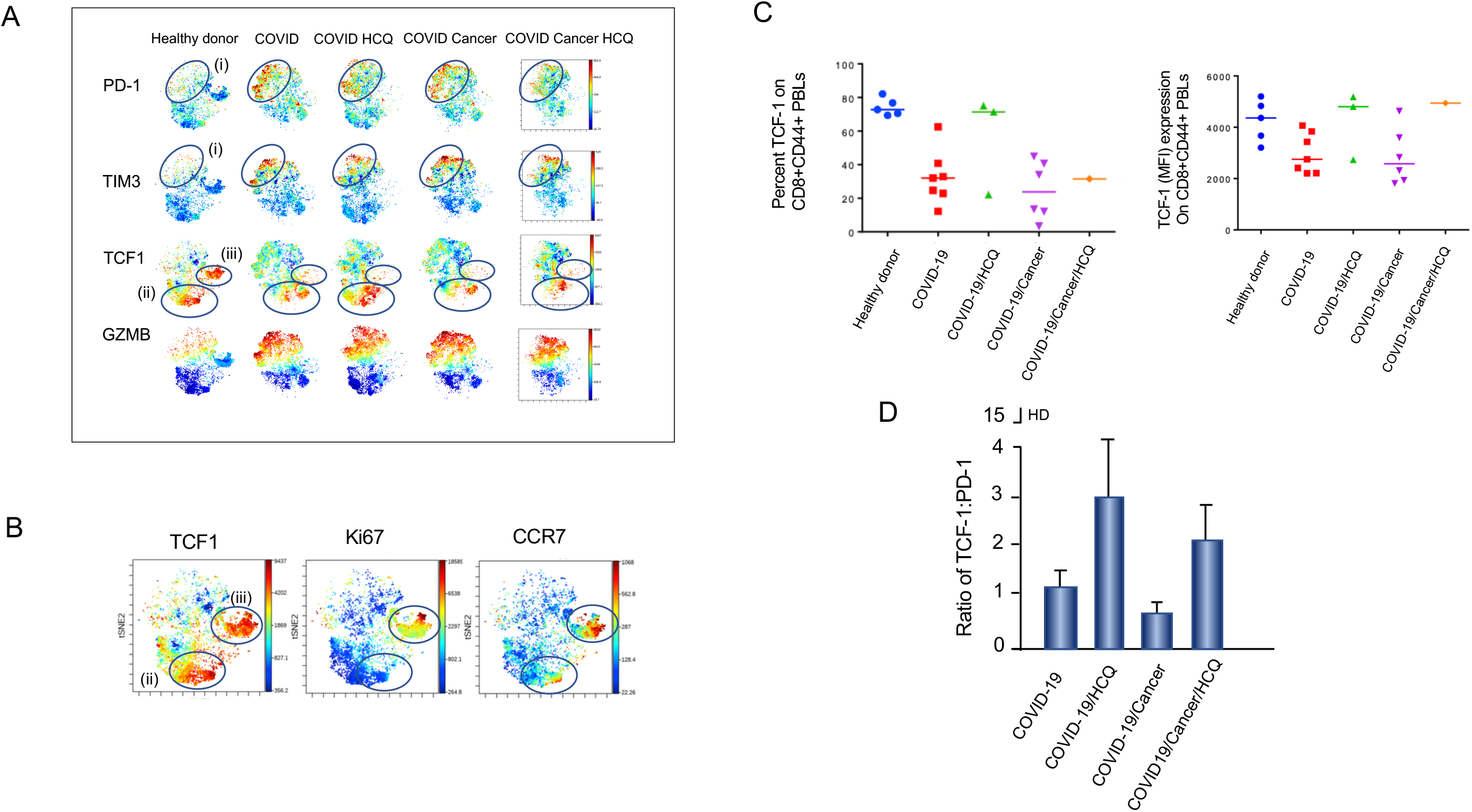
TCF-1 is a marker in peripheral blood T-cells for responses to SARs CoV2 and HCQ. **Panel A**: viSNE patterns of murine peripheral T-cells stained with anti-PD-1, TIM-3, TCF-1 and GZMB. **Panel B:** viSNE patterns of TCF-1 expression (left panel) as well as Ki67 (middle panel) and CCR7 (right panel). **Panel C:** Dot plot analysis showing the MFI of TCF-1 expression showing that Covid-19 reduces the expression of TCF-1 on CD8+ peripheral T-cells. Treatment with HCQ inhibited this decrease. c **Left panel:** Dot plot analysis showing the percentage of TCF-1 expressing CD8+CD44+ peripheral T-cells. **Right panel:** Dot plot analysis showing the MFI of TCF-1 expressing CD8+CD44+ peripheral T-cells. **Panel D:** Ratio of TCF-1 to PD-1

CD8+ T-cells from the blood of COVID-19 patients relative to healthy donors showed an increase in the staining with anti-PD-1, TIM-3 and GZMB, concurrent with a reduction in the staining of both TCF-1+ islands (**Fig. 6B**). By contrast, HCQ treatment increased the presence of TCF-1 and CD8+TCF-1+Ki67-CCR7+/-cells in island (ii), both in the samples from patients with the COVID-19. HCQ treatment had no effect in the CD8+TCF-1+Ki67+CCR7+ (iii) subset. The same pattern was seen in COVID-19 patients with cancer, and in the one sample we were able to obtain from a patient treated with COVID-19/cancer/HCQ. All cancer patients had either pancreatic or oesophageal cancer who had been treated with chemotherapy. These data showed that HCQ could prevent the loss of TCF-1+ progenitors in blood with a less activate or differentiated phenotype.

SARs-CoV-2 infection decreased the overall expression of TCF-1 staining both in terms of the percentage of cells and the intensity of staining (MFI **(Fig. 6C**) and in the ratio of TCF-1 to PD-1 MFIs (**Fig. 6D**). HCQ showed a trend in reversing this decrease and in the ratio of TCF-1 to PD-1. Overall, our findings showed that the expression of TCF-1 could be used as a marker for the both SARs CoV2 and treatment with HCQ.

## Discussion

Globally, HCQ continues to be used to treat thousands of COVID-19 patients with new and ongoing clinical trials underway to establish its efficacy. This situation makes an inquiry into the effects on HCQ and AZ on cancer immunotherapy an immediate pressing issue. In this study, we show that HCQ alone, or in combination with AZ, at the doses used to treat COVID-19 patients, reverses the ability of anti-PD-1 therapy to control the growth of cancer. This reversal was accompanied by a selective interference with CD8+TCF1+ progenitor and CD8+PD-1+ effector TIL expansion or infiltration of tumors. Further, we show that TCF-1 expression serves as a marker in peripheral blood CD8+ T-cells for the response of the immune system to SARs-CoV-2 and HCQ therapy. The reduction in the presence of CD8+ subsets would account for the diminished anti-tumorigenic efficacy of PD-1 immunotherapy. Our study underscores the need for caution in the use of HCQ in the treatment of COVID-19 patients under immunotherapy.

The administration of HCQ, either alone, or together with AZ, at doses used in COVID-19 studies, impaired the ability of anti-PD-1 therapy to reduce the growth of B16 melanoma tumors in mice. The reversal of the benefits was seen as early as days 12-16 post-implantation, while no effect was observed on the growth of the B16 tumor in mice not treated with immunotherapy. The effects were seen on the composition of infiltrating CD8+ T-cells and not on the growth of the tumor cells themselves. Mechanistically, HCQ and HCQ/AZ reduced an anti-PD-1 induced increase in the expression of PD-L1 on tumor cells, while exerting specific inhibitory effects on the presence of progenitor and effector CD8+ TILs. While the reduction in PD-L1 expression of B16 cells should have promoted tumor rejection, the concurrent inhibition of CD8+ progenitors and effectors appeared to dominant in the inhibition and reversal of the anti-PD-1 anti-tumor response. The targeting of CD8+ T-cells by HCQ in anti-PD-1 treatment is consistent with the fact that CD8+ T-cells are the prime target of immunotherapy elicited by anti-PD-1. We found no statistically significant effect on the presence of CD4+ T-cells, B-cells or myeloid cells. In this context, it is also noteworthy that T- and B-cells use different signaling pathways to initiate proliferation (Rudd, 1999; Wilkinson et al., 2004). Differences in signaling have also been noted between CD4 and CD8+ T-cells where IL-2 induces quantitatively stronger proliferation in CD8^+^ T cells compared to CD4^+^ T cells (Gesbert et al., 2005) due to greater IL-2Rβ chain expression (Smith et al., 2017). We also failed to find differences in antibody titres in mice or in levels of IgG1/G2 antibodies against PD-L1. This susceptibility of CD8+ T-cells to inhibition by HCQ in the context of anti-PD-1 immunotherapy may also involve factors in the tumor microenvironment that affected immune activation and function.

As reported by others (Siddiqui et al., 2019; Wei et al., 2017), we observed that anti-PD-1 immunotherapy increased the presence of PD-1+ TCF-1+ progenitors and PD-1+CD44+TCF-1-effector TILs. These progenitors in turn are needed to produce more differentiated PD-1+TCF1-cells in immunotherapy (Siddiqui et al., 2019). Importantly, HCQ and HCQ/AZ inhibited the ability of anti-PD-1 therapy to increase in the presence of these PD-1+TCF-1+ progenitors. Consistent with this, HCQ also inhibited the ability of anti-PD-1 to expand CD8+CD44+ TILs. These cells generally expressed low-intermediate levels of the activation markers PD-1 and TIM-3, and most likely represent a newly expanded effector population expressing these activation markers. However, co-expression of PD-1 and TIM-3 with TOX may also correspond to an early stage of cellular expansion leading to T-cell exhaustion. In our model, anti-PD-1 therapy reduced the presence of CD8+ TILs with the high co-expression of PD-1, TIM-3 and TOX indicative of terminal exhaustion (Anderson et al., 2016; McLane et al., 2019). Our study was able to distinguish between subsets with different levels of receptor expression showing the anti-PD-1 preferentially expands CD8+ effector cells with low-intermediate levels of PD-1, TIM-3 expression and actually limits the presence of more terminally exhausted CD8+ cells. Differences in these findings could be related to the tumor type and the duration in anti-PD-1 treatment of mice. HCQ concurrently inhibited the anti-PD-1 induced reduction in this subset of terminally exhausted T-cells, although the effect of the combination of HCQ/AZ did not inhibit the reduction, in the case of the CD8+ cells with highest observed levels of PD-1, TIM3 and TOX indicative of the most severe exhaustion. Our observations are most consistent with a general ability of HCQ to inhibit the activation and proliferation of CD8+ TILs. It can inhibit the appearance of CD8+ progenitors, newly generated CD8+ effectors from progenitors and the progression CD8+ effectors to a more exhausted phenotype.

Surprisingly, AZ alone failed to elicit a statistically significant effect in mice on tumor growth as observed for HCQ. AZT and ciprofloxacin (CPX) act as lipophilic weak bases where they affect intracellular organelles similar to CQ (Poschet et al., 2020) and previous studies have documented a role for antibiotics such as ciprofloxacin (CPX) in the reversal of immune checkpoint blockade on tumour eradication (Routy et al., 2018; Vetizou et al., 2015). These previous studies emphasized effects on the gut microbiota (Routy et al., 2018; Vetizou et al., 2015). The differences in these findings may relate to the dose of antibiotics used or the coverage of bacterial inhibition by AZT versus ciprofloxacin (CPX) and other antibiotics. It might also relate to the nature of gut biota in mice which is likely to vary in different containment facilities.

In terms of the expression of specific molecules, HCQ had variable effects on markers indicative of activation. For example, while HCQ inhibited the expression of TCF1 TILs, it did not reproducibly affect the expression of the activation marker Ki67 in progenitor T-cells. One surprising observation was the potent effect of HCQ in increasing CD8 expression on progenitor and effector T-cells. It is paradoxical given our previous findings showing that CD8 (and CD4) bind to p56^lck^ to initiate a protein-tyrosine phosphorylation cascade in T-cells that is needed for T-cell activation (Barber et al., 1989; Rudd et al., 1988). Further, the down-regulation of CD8-lck has been suggested as a mechanism for peripheral tolerance (Schonrich et al., 1991; Zhang et al., 1995), while subpopulations with low CD8 have been reported during chronic diseases (Grisotto et al., 2001) and in acute immune responses to pathogens (Walker et al., 1995). HCQ and AZ increased CD8 expression in combination with anti-PD-1. No effect was seen in the absence of anti-PD-1 indicating that altered signaling linked to anti-PD-1 blockade synergizes with HCQ and AZ to promote increased CD8 expression. It was observed on progenitors and effector T-cells. The functional consequences of this increased expression are unclear, although an increased number of CD8 molecules engaged by MHC during antigen-presentation would be expected to increase the activation and proliferation of T-cells (Rudd, 1999). Alternatively, depending on the relative stoichiometry and localisation of expression, an excess of CD8 receptors might lead to few receptors bound to p56^lck^ and hence, reduced activation. CD4 sequestration of p56^lck^ complex may also prohibit the induction of activation signals through the TCR-CD3 complex (Haughn et al., 1992).

Lastly, we were able to identify TCF-1+ expression on peripheral T-cells as a marker for the effects of HCQ and SARs-CoV-2 infection. viSNE analysis identified a population of TCF-1+ T-cells which was distinct from other effector CD8+ T-cells expressing GZMB/PD-1/TIM-3. TCF-1 expression was markedly reduced in the PBLs from patients infected with SAR-CoV-2 infection which conversely was associated with a less prominent increase in GZMB/PD-1/TIM-3 expressing effector T-cells. HCQ therapy blocked this decrease in TCF-1 expression, both in terms of the MFI and the percentage of CD8+ cells expressing the transcription factor. A similar effect was seen in the ratio of TCF-1 versus PD-1 expression. Changes in TCF-1 expression might be useful as a surrogate marker for SARs-CoV-2 infection and the treatment of patients with HCQ.

Many questions remain related to HCQ, COVID-19 and tumor immunology. Whether the inhibitory effects on anti-PD-1 immune therapy may be offset by its reported benefit in sensitizing chemotherapy remains to be determined. For example, HCQ has been reported to enhance the anti-cancer activity of the histone deacetylase inhibitor, vorinostat (VOR), in pre-clinical models and early phase clinical studies of metastatic colorectal cancer (mCRC) (Patel et al., 2016). Lastly, the inhibitory effects on anti-PD-1 therapy in this study used HCQ doses that have been used in the treatment of COVID-19 patients (Chen et al., 2020a; Chen et al., 2020b; Gao et al., 2020; Gautret et al., 2020; Geleris et al., 2020; Magagnoli et al., 2020; Mahevas et al., 2020; Molina et al., 2020; Rosenberg et al., 2020; Tang et al., 2020; Yu et al., 2020). It remains unclear whether lower concentrations of drug will have the same effect, although the effectiveness of lower concentrations in treating auto-immune diseases such as SLE is in part likely due to its immune-suppressive properties (Thome et al., 2014; Thome et al., 2013). The utility of HCQ may vary dependent on the dose of the drug and the nature of cancer therapy.

## Materials and Methods

### Housing and ethical approval

C57BL/6 mice were housed at the Hôpital Maisonneuve Rosemont animal facility (Montreal, QC, Canada). Mice were housed in individually ventilated cages (IVC) and all experiments were approved by the CR-HMR Ethical Approval (Le Comité de protection des animaux du CIUSSS de l’Est-de-l’ILle-de-Montréal (CPA-CEMTL), F06 CPA-21061 du projet 2017-1346, 2017-JA-001). The human study was approved by the McGill University Health Centre (MUHC) Research Ethics Board #2021-6881. The peripheral blood lymphocytes were isolated at the MUHC-RI by MUHC staff using the density gradient centrifugation method. Cells were washed 3 times in PBS and fixed in 3.5% paraformaldehyde at room temperature for 30 minutes for the inactivation of the SARS coronavirus as described (Kariwa et al., 2004). COVID-19 patients were treated with 400mg HCQ twice a day (BID) on day 1 followed by 200 po BID for the next 5 days, reassessed based on progression (BID = 2x daily). Patients were monitored for hemolytic anemia with CBC every 2 days. Testing for G6PD levels was carried out on patients considered at higher risk based on ethnicity, hemolytic anemia and family history (Empiric Management of Suspected COVID-19 cases (adults) (MUHC).

### Mice

C57BL/6 mice were housed at the Hôpital Maisonneuve Rosemont animal facility (Montreal, QC, Canada). Mice, aged 7-8 weeks, were implanted intradermally with 50,000 B16-PD-L1 melanoma cells that overexpress PD-L1. Cells were kindly provided by Dr. M. Ardolino, Ottawa, Canada. Anti-PD-1 treatment (200ug/mouse, i.p., 2x/week) started at day 4 post-implantation, when tumors were established. After 1-week of anti-PD-1 treatment, first signs of tumor response were noted and HCQ (40 mg/kg, daily) and/or AZ (40 mg/kg for day 1 and 2; 20 mg/kg for day 3-5) were injected i.p. When giving combined, injections were performed within a 4 h time interval. Due to different pharmacokinetics of HCQ in mice, 40 mg/kg in mouse represents 600 mg in humans (Jackson et al., 2016, abstract, AACR). Tissues and PBMCs were harvested day 17 post-implantation.

### Chemicals and antibodies

Hydroxychloroquine sulfate (HCQ) was purchased from Selleckchem and azithromycin (AZ) was purchased from Hemopharm.

### Mouse tissue collection and preparation

Tumour infiltrating lymphocyte collection: Briefly, tumors were harvested and minced into small pieces before enzymatic digestion with Liberase TL (Sigma, Damstadt, Germany) and DNase1 (Thermo Fisher, Waltham, MA, USA) at 37°C for 30 min. Addition of RPMI with 10% FBS stopped reaction and mixture were passed through 70 µm cell strainers to generate single cell suspension. After 2 washes cells were (i) for total TILs quantification stained for viability and fixed or (ii) overlayed on a ficoll layer for lymphocyte collection. Lymphocytes were then stained for viability followed by surface staining with FACS antibodies before fixation. Intracellular staining was performed using the FoxP3/Transcription Factor Staining Buffer Set (ebiosciences, San Diego, CA, USA). For PBL isolation from mice, blood was collected in EDTA-coated tubes and peripheral blood mononuclear cells (PBMCs) were isolated by Ficoll (Corning, NY, USA). After 3 washes, PBMCs were stained for FACS. For the preparation of thymocytes, thymi were collected from all mice immediately after sacrifice, dissociated and passed through 70 µm cell strainers into RPMI medium. Following 3 washes, cells were stained for viability and fixed in 2% PFA until further analysis.

### Tumour infiltrating lymphocyte collection

Briefly, tumours were dissected and placed in RPMI medium on ice. Tumours were minced with razor blades into pieces and then digested in the presence of liberase and DNase at 37°C for 30 min, with manual shaking every 5 min. The mixture was then strained through µm cell strainers, and collected cells were rinsed twice in 2 % FCS-PBS and overlayed over on a fill layer for lymphocyte. After ficoll separation, TILs were collected and subjected to viability staining, then fixed with 2% PFA.

### Flow Cytometry and antibodies

Antibodies were purchased from either Biolegend or BD Biosciences. Flow cytometric analyzes were performed according to standard protocols. In brief, PFA-fixed TILs were washed twice with cold PBS and stained with the targets listed below for 30 min on ice and then permeabilized for 45 min on ice. TILs were then stained for intracellular targets for 45 min on ice, and then washed twice with PBS and proceeded to data collection. The antibodies used as listed below:

| Stain | Fluorophore | Manufacturer |
| --- | --- | --- |
| Viability | BV510 | BD Biosciences |
| CD45 | AF700 | Biolegend |
| TCRb | BUV395 | BD Biosciences |
| CD19 | PE | Biolegend |
| CD11c | BV650 | Biolegend |
| CD11b | APC | Biolegend |
| PD-L1 | PerCP/Cy5.5 | Biolegend |
| CD31 | PECy7 | BD Biosciences |
| Ly6C | BV605 | Biolegend |
| Ly6G | APCCy7 | Biolegend |
| MHCII | BV421 | BD Biosciences |
| PD-1 | BV786 | Biolegend |
| CD80 | FITC | Biolegend |

Intracellular stainings were performed using the FoxP3/Transcription Factor Staining Buffer Set (ebiosciences, San Diego, CA, USA). Cells were acquired using BD LSR Fortessa X-20 and DIVA software. FACS analyses were performed using FlowJo software and Cytobank premium. The antibodies were used to stain subsets of immune cells as outlined in the table below:

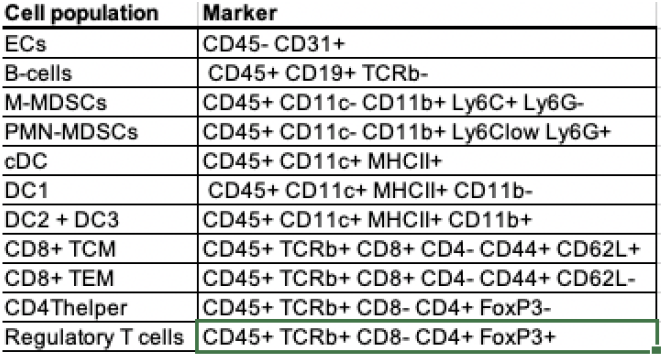

### Human PBMC isolation and FACs staining

Blood was collected from healthy donor, cancer or COVID-19 patients (MUHC, Montreal, Canada). Blood was collected from healthy donor, cancer or COVID-19 patients. Peripheral blood mononuclear cells (PBMCs) were freshly isolated by dextran sedimentation (Corning, Corning, NY). Then, cells were fixed with a 3% paraformaldehyde solution during 30min at room temperature. Finally, cells were frozen in FBS 7.5% DMSO and storage at -80°C until the FACs staining. The day of the FACs staining, PBMC were thawed and washed in PBS 2% FBS (FACs buffer). PBMC were stained with PerCP Cy5.5-conjugated anti-CD3 (clone: UCHT1), BV711-conjugated anti-CD8 (clone: RPA-T8), BV605-conjugated anti-PD-1 (clone: EH12.1) and APC-Cy7-conjugated anti-CD69 (clone: FN50) in FACs buffer during 20min at 4°C in the dark. After, cells were washed and fixed in Fixation/Permeabilization Buffer during 40min at 4°C in the dark and finally stained with PECF594-conjugated anti-Granzyme B (clone: GB-11) during 30min in Permeabilization Buffer. The acquisition was performed by using BD LSRFortessa X-20 and DIVA software (Beckton Dickinson). FACS analyses were performed by using FlowJo software or Cytobank for viSNE analyses.

### Wholemount brain staining

Brains were fixed in 4% paraformaldehyde overnight at 4°C, washed 3 times in PBS, and blocked overnight in blocking solution (0.1Tris-HCl, 150 mM NaCl, 0.2% Blocking reagent (PerkinElmer), 0.5% Triton-X). After overnight incubation with anti-Smooth Muscle Actin-FITC (Sigma), brains were washed and imaged with the use of a fluorescent dissecting microscope.

### Retinal staining

Eyes were prefixed in 4% paraformaldehyde for 15 min at room temperature. Dissected retinas were blocked overnight at 4° C in blocking solution (0.1Tris-HCl, 150 mM NaCl, 0.2% Blocking reagent (PerkinElmer), 0.5% Triton-X). Retinas were then incubated with IsolectinB4 and immunostained with anti-Smooth Muscle Actin-FITC (Sigma).Quantification of retinal vasculature branch points was done with the use of the angiogenesis analyzer tool for ImageJ.

### Wholemount brain and retinal staining

Brains were fixed in 4% PFA overnight at 4°C, washed 3 times in PBS, and blocked overnight in blocking solution (0.1Tris-HCl, 150 mM NaCl, 0.2% Perkin Elmer blocking reagent and 0.5% Triton-X). After overnight incubation with anti-Smooth Muscle Actin-FITC (Sigma), brains were washed and imaged with the use of a fluorescent dissecting microscope.

Eyes were prefixed in 4% PFA for 15 min at room temperature. Dissected retinas were blocked overnight at 4°C in blocking solution (0.1Tris-HCl, 150 mM NaCl, 0.2% Perkin Elmer blocking reagent and, and 0.5% Triton-X). Retinas were then incubated with IsolectinB4 and immunostained with anti-Smooth Muscle Actin-FITC (Sigma). Quantification of retinal vasculature branch points was done using the angiogenesis analyzer tool for ImageJ.

### Anti-B16-PDL1 antibody assay

Plasma was collected from heparinized murine peripheral blood by cetrifugation for 15 min at 2000g. Samples were immediately used or aliquoted and frozen at -80°C. For the antibody assay we used a slightly modified version of Dilillo D. et al. 2012 protocol. Briefly, 5×10^5^ B16-PDL1 tumour cells were added to a 96-well plated, blocked with 10% FBS-PBS, and washed with PBS. Plasma samples were diluted 1:16 in PBS and were added to B16 cells in a volume of 100 μl, mixed and then incubated for 1 hour at 4°C. Cells were intensively washed and secondary Goat anti-mouse IgG-af488 conjugated (A11029; Invitrogen) and Rat anti-mouse IgM-APC (clone RMM-1; Biolgend #406509) both at 1:500 dilution and incubated on ice for additional 1 hour. Cells were washed and passed to flow cytometry. Figure S5. Shows data from one experiment that is representative of all experiments held. Median fluorescence intensity (MFI) and the frequency of positively stained cells were assessed in FlowJo (Treestar). Index value was calculated by normalizing B16-IgG and B16-IgM MFI of the sample to the average of tumor-free mice.

### Statistics

All data are expressed as mean ± SEM. A t-test was used when only two groups were compared, and a one-way ANOVA was used when more than two groups were compared. A two-way ANOVA was used for multiple comparison procedures that involved two independent variables. Dunett or Bonferroni tests were used for *posthoc* analyses. A difference in mean values between groups was significant when p ≤ 0.05 *; p ≤ 0.01 ** and p ≤ 0.001 ***.

## Data Availability

All data referred to in the paper is available.

## SUPPLEMENTAL INFORMATION

Supplemental Information can be found online at xxxxxx

## ACKNOWLEDGMENTS

C.E.R. was supported by Canadian Institutes of Health Foundation grant (159912).

## AUTHOR CONTRIBUTIONS

J.K, F.S., M.I. and A.K. and C.E.R. designed the study. J.K, F.S., M.I. and A.K. conducted most experiments. R.K and C.M. facilitated and arranged for the identification of clinical samples as well as arranging for ethical approval for their use. C.E.R. wrote the manuscript with assistance from J.K, F.S., M.I.,R.K., C.M. and A.K.

